# Influenza H5N1 and H1N1 viruses remain infectious in unpasteurized milk on milking machinery surfaces

**DOI:** 10.1101/2024.05.22.24307745

**Authors:** Valerie Le Sage, A.J. Campbell, Douglas S. Reed, W. Paul Duprex, Seema S. Lakdawala

## Abstract

Spillover of highly pathogenic avian H5N1 into the cattle population poses a risk to humans through the close contact with farm workers. High viral loads of influenza viruses in the unpasteurized milk of infected lactating cows has the potential to contaminate equipment within milking parlors and create fomites for transmission to dairy workers. Cattle H5N1 and human 2009 H1N1 pandemic influenza viruses were found to remain infectious on surfaces commonly found in milking equipment materials for a few hours. The data presented here provide a compelling case for the risk of contaminated surfaces generated during milking to facilitate transmission of H5N1 from cattle-to-cattle and to dairy farm workers.

---

Highly pathogenic avian influenza virus was first detected in US domestic dairy cattle at the end of March 2024, and has since spread to herds across multiple state lines and resulted in at least one confirmed human infection [1]. Assessment of milk from the infected dairy cows indicated that unpasteurized milk contained high levels of infectious virus [2]. Exposure of dairy farm workers to contaminated unpasteurized milk during the milking process poses a large threat to humans that may lead to 1) increased H5 infections in dairy farm workers and 2) the possibility for H5 viruses to adapt within humans and gain the capability of human-to-human transmission.

The milking process is primarily automated with machines (commonly referred to as ‘claws’) that attach to the dairy cow udder for milk collection (Figure 1A). However, a number of steps require human input in the process, including forestripping, whereby the first 3-5 streams of milk are expressed from each teat by hand. This process stimulates the teats for optimal milk release, improves milk quality by removing bacteria, and provides an opportunity to check for abnormal milk. The forestripping process can result in milk splatter on the equipment as well as the production of aerosols into the milking parlor from cows that are releasing milk. The udders are cleaned and dried by hand prior to installation of the claw onto each teat. Housed within a stainless steel shell, the flexible rubber inflation liner makes contact with the teat and expands and contracts to release the milk (Figure 1A). The claw apparatus automatically adjusts to the flow rate of the milk, and once the flow rate has slowed, the inflation liners remove the pressure on the udders and the machine releases from the animal. This release can also result in milk spray onto dairy farm workers, the equipment, or the surrounding areas, and the possible formation of aerosols into the environment. Importantly, milking often takes place at human eye level, with the human workspace physically lower than the cows, which increases the potential for contact of infectious milk with mucus membranes. Currently, no eye or respiratory protection is required for dairy farm workers.

**Figure 1:**
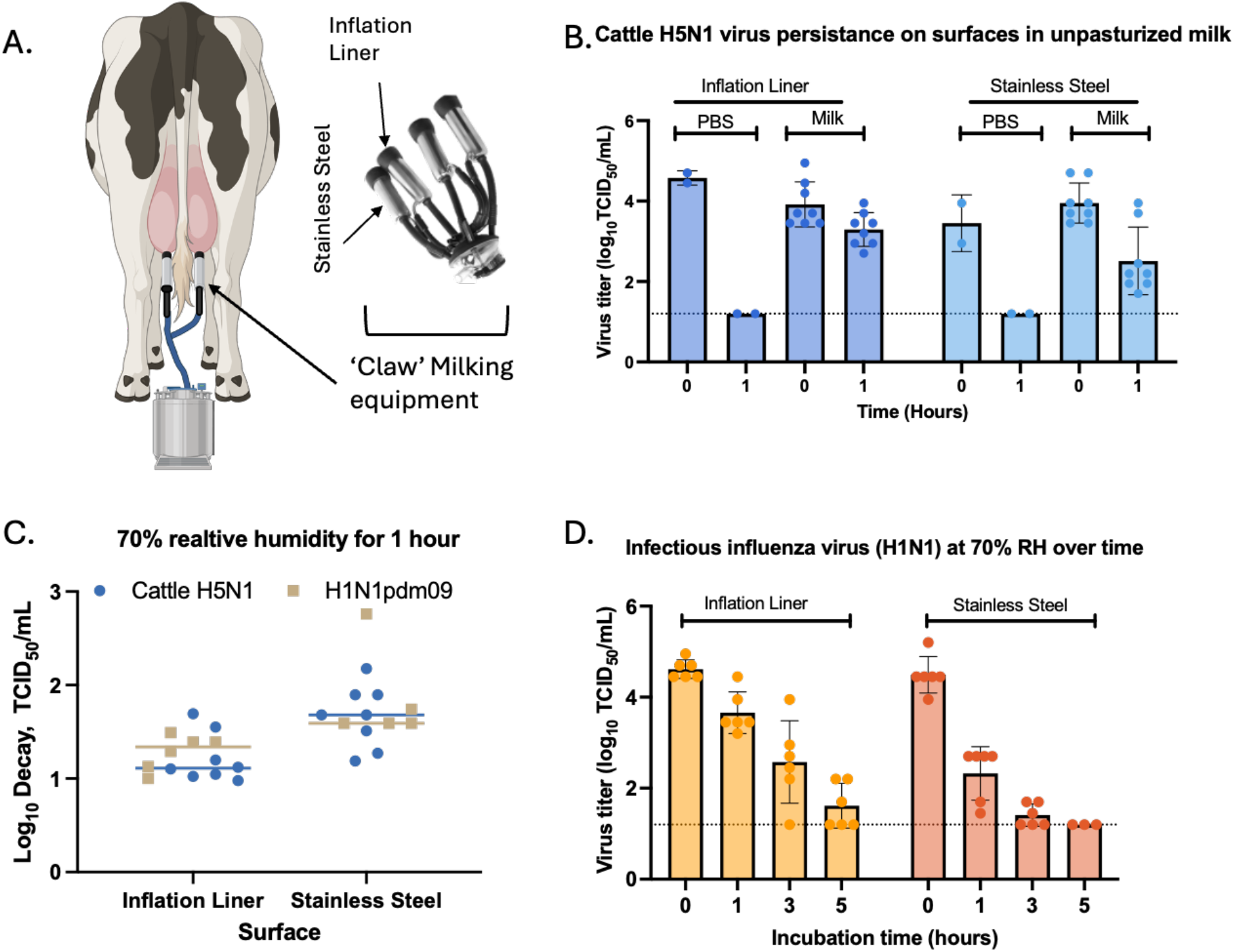
Persistence of influenza virus in unpasteurized milk on inflation rubber liner and stainless steel. A) Schematic of milking equipment and the materials tested for virus stability. Created in Biorender.com B) Viral titers of the cattle H5N1 virus diluted in unpasteurized milk and deposited as 10 1uL droplets onto the indicated surfaces. Droplets were recovered immediately post deposition (time 0) or 1 hour after aging at 70% relative humidity (RH). C) Comparison of log decay values of H5N1 and H1N1 virus in unpasteurized milk at 70% relative humidity for 1 hour on the two indicated materials. Log decay was calculated as a ratio of the viral titer at time 0 divided by the titer after 1 hour of aging. D) Viral titers of the H1N1 virus in unpasteurized milk on the two surfaces at 70% relative humidity for 0, 1, 3, or 5 hours of aging. In B-D, each symbol is an individual replicate of at least two biological replicates using two distinct lots of unpasteurized milk performed in technical triplicates. Virus titer was calculated by determining the traditional tissue culture infectious dose 50 (TCID_50_) assay on Madin-Darby canine kidney cells. All raw data can be accessed at: https://doi.org/10.6084/m9.figshare.c.7242034.v1.

The persistence of influenza viruses in unpasteurized milk on surfaces is unknown, but this information is critical to understand the viral exposure risk to dairy workers during the milking process. We analyzed the persistence of infectious H1N1 and H5N1 influenza viruses in unpasteurized milk on surfaces commonly found in milking equipment, such as inflation rubber liners and stainless steel (Figure 1A). Influenza viruses, A/dairy cattle/TX/8749001/2024 (H5N1) or a surrogate virus, the 2009 pandemic strain A/California/07/2009 (H1N1), were diluted in raw unpasteurized milk. Similar to prior studies [3-5], small droplets of diluted virus were pipetted onto either stainless steel or inflation rubber liner coupons within an environmental chamber and collected immediately (time 0), or after 1, 3 or 5 hours of aging. To mimic environmental conditions within open-air milking parlors in the Texas panhandle in March and April 2024, persistence studies were conducted using 70% relative humidity (RH).

Importantly, we observed that the H5N1 cattle virus remained infectious in unpasteurized milk on stainless steel and inflation lining rubber compared to PBS (phosphate buffered saline) over 1 hour (Figure 1B). This indicates that unpasteurized milk containing the H5N1 virus will remain infectious on milking equipment. To assess whether a less pathogenic influenza virus could be used as a surrogate to study important aspects of viral persistence, we compared viral decay between H5N1 and H1N1 in raw milk over 1 hour on these two surfaces (Figure 1C). The similar decay of these two viruses on both surfaces suggests that H1N1 can be used as a surrogate for H5N1 cattle virus in studies regarding viral persistence in raw milk. Further experiments examining the infectiousness of H1N1 over a longer period of time revealed the persistence on inflation liner for at least 3 hours and for at least 1 hour on stainless steel when in unpasteurized milk (Figure 1D). These results indicate that influenza virus is very stable in unpasteurized milk and that deposited H5N1 on milking equipment could remain infectious for long periods of time.

Taken together, the data presented here provide a compelling case for the risk of contaminated surfaces generated during milking to infect dairy farm workers with H5N1 virus. Implementation of personal protective equipment such as face shields, masks, and eye protection are needed to ensure reduced spillover of H5N1 from dairy cows to humans. In addition, contaminated inflation liners or other milking equipment may be responsible for some of the cattle-to-cattle spread observed on dairy farms. The use of disposable liners or sanitization of the liners between cows could assist in reducing the spread of influenza virus between the animals at a facility to curb the current outbreak.

## Data Availability

All raw data can be accessed at: https://doi.org/10.6084/m9.figshare.c.7242034.v1.

## Acknowledgements

This project has been funded in part with Federal funds from the National Institute of Allergy and Infectious Diseases, National Institutes of Health, Department of Health and Human Services, under Contract No. 75N93021C00015 and an NIH award (UC7AI180311) from the National Institute of Allergy and Infectious Diseases (NIAID) supporting the Operations of The University of Pittsburgh Regional Biocontainment Laboratory (RBL) within the Center for Vaccine Research (CVR). We thank the Lakdawala lab members, CEIRR risk assessment pipeline meeting attendees, Rachel Duron, and Linsey Marr for useful feedback.

